# SARS-CoV-2 Antibody Prevalence and Association with Routine Laboratory Values in a Life Insurance Applicant Population

**DOI:** 10.1101/2020.09.09.20191296

**Authors:** Steven J. Rigatti, Robert Stout

## Abstract

**Objectives:** The prevalence of SARS-CoV-2 antibodies in the general population is largely unknown. Since many infections, even among the elderly and other vulnerable populations, are asymptomatic, the prevalence of antibodies could help determine how far along the path to herd immunity the general population has progressed. Also, in order to clarify the clinical manifestations of current or recent past COVID-19 illness, it may be useful to determine if there are any common alterations in routine clinical laboratory values.

**Methods:** We performed SARS-CoV-2 antibody tests on 50,130 consecutive life insurance applicants who were having blood drawn for the purpose of underwriting (life risk assessment). Subjects were also tested for lipids, liver function tests, renal function studies, as well as serum proteins. Other variables included height, weight, blood pressure at the time of the blood draw, and history of common chronic diseases (hypertension, heart disease, diabetes, and cancer).

**Results:** The overall prevalence of SARS-CoV-2 was 3.0%, and was fairly consistent across the age range and similar in males and females. Several of the routine laboratory tests obtained were significantly different in antibody-positive vs. antibody-negative subjects, including albumin, globulins, bilirubin, and the urine albumin:creatinine ratio. The BMI was also significantly higher in the antibody-positive group. Geographical distribution revealed a very high level of positivity in the state of New York compared to all other areas (17.1%). Using state population data from the US Census, it is estimated that this level of seropositivity would correspond to 6.98 million (99% CI: 6.56-7.38 million) SARS-CoV-2 infections in the US, which is 3.8 times the cumulative number of cases in the US reported to the CDC as of June 1, 2020.

**Conclusions:** The estimated number of total SARS-CoV-2 infections based on positive serology is substantially higher than the total number of cases reported to the CDC. Certain laboratory values, particularly serum protein levels, are associated with positive serology, though these associations are not likely to be clinically meaningful.

## Introduction

In early 2020 a novel coronavirus emerged in Hubei Province, China^1^. The causative agent was a betacoronavirus most closely related genetically to zoonotic viruses found in bats, and clinically similar to recent emergent epidemic coronaviruses which caused Severe Acute Respiratory Syndrome (SARS) and Middle Eastern Respiratory Syndrome (MERS)^2^. Since then, the virus has become a worldwide pandemic, infecting over 17 million persons and causing more than 660,000 deaths as of this writing^3^. The first case in the United States occurred on January 20^th^, 2020^4^. And since then the Centers for Disease Control and Prevention has recommended that all states report laboratory-confirmed cases^5^. Case counts have been closely tracked by the CDC, the press, and academic institutions. However, because the illness caused by SARS-CoV-2 may be asymptomatic or minimally symptomatic^6^, these counts of cases may underestimate the number of persons who have been infected. Various studies of seroprevalence in the United States^7,8^ have shown different results based on timing and locality, but have been consistent in showing that seroprevalence is higher than would be implied by simple case counts based on viral antigen testing.

Because SARS-CoV-2 is novel, the presence of antibodies in the blood likely indicates a history of infection since the pandemic began, and serologic testing can be used to estimate the overall rate of infection, even in those who had minimal symptoms or who were never tested despite symptoms.

In this study, a convenience sample of blood specimens submitted to a commercial laboratory was used to conduct a survey of seroprevalence. The goal was both to estimate the overall number of cases in the general population and to examine the data to determine if any common clinical laboratory tests were significantly associated with seropositivity.

## Methods

In the United States, the process of purchasing life insurance often involves a brief physical examination by paramedical professionals, the collection of height, weight and blood pressure measurements, and the testing of blood and urine specimens for common analytes related to overall health. Such tests are seldom, if ever, performed on individuals below age 17 years or above 85 years. Also, blood tests are generally reserved for individuals applying for higher dollar amounts of life insurance or for those applying for permanent types of insurance (rather than term insurance). Thus, individuals applying for life insurance are a self-selected group primarily from higher socio-economic strata. Those who have a history of chronic illness may be less likely to apply because more serious conditions can be associated with higher life premiums.

Between May12^th^ and June 25^th^ 2020, 50,130 individuals were tested for antibodies to SARSCoV-2. Individuals were part of a convenience sample from a pool of life insurance applicants who had blood tests performed as part of life insurance underwriting at Clinical Reference Laboratories. This sample represents approximately one fifth of all samples tested at the facility during that time. All applicants self-reported that they were well at the time of application. The antibody tests were performed using the Roche Elecsys Anti-SARS-CoV-2 kit on the Roche 602 analyzer, with a stated sensitivity of 100% and specificity of 99.8%, utilizing an electrochemiluminescence immunoassay.

Western Institutional Review Board’s (WIRB’s) IRB Affairs Department reviewed the study under the Common Rule and applicable guidance and determined it is exempt under 45 CFR §46.104(d)(4) using de-identified study samples for epidemiologic investigation. within one year), height, weight, blood pressure, and routine laboratory measures which analyzer, with a stated sensitivity of 100% and specificity of 99.8%, utilizing an electrochemiluminescence immunoassay.

Other information available on test subjects included age, sex, smoking status (tobacco use within one year), height, weight, blood pressure, and routine laboratory measures which included some combination of glucose, blood urea nitrogen, creatinine, aspartate aminotransferase, alanine aminotransferase, gamma glutamyltransferase, alkaline phosphatase, total bilirubin, total cholesterol, high density lipoprotein (HDL) cholesterol, triglycerides, lactate dehydrogenase, hemoglobin A1c, and NT-pro B-type natriuretic peptide.

Limited medical history was available in the form of responses to simple yes/no questions regarding a history of cardiovascular disease, diabetes, hypertension, and cancer.

The differences in continuous variables between the antibody-positive and negative groups were tested for significance with the Mann-Whitney U test, while differences in categorical variables were tested using the chi-square test.

To estimate the total burden of SARS-CoV-2 infections in the US, census data was obtained. For each state and the District of Columbia, the total 2018 estimated census population was multiplied by the US population proportion between the ages of 20 and 80 (71.1%). Then, the state-specific proportion of positive tests was applied from our sample. Confidence limits were estimated by generating 5000 bootstrap samples (with replacement) of our data and recalculating the total number of US cases. Under and over-representation of states was determined by a ratio between the proportion of individuals living in a given state to the proportion of tests performed in that state.

All statistical analyses were performed using R (version 3.6.1)^9^ and R-studio (version 1.2.1335)^10^.

## Results

The overall sample included 50,025 individuals with a median age of 42 years (IQR: 34-54), 56% of whom were male. Geographical distribution deviated somewhat from the overall population distribution of the US, with some under-representation from Maine, West Virginia, Vermont and Oklahoma, and over-representation from Nebraska, Hawaii, and Utah. Characteristics of the study population are displayed in Table 1. The antibody positive group tended to be slightly younger (median age 41) vs. the antibody negative group (median age 42). The proportion of subjects reporting a history of heart disease, hypertension and/or diabetes was similar between the positive and negative groups. Laboratory tests which were statistically different (p < 0.01) between the positive and negative group included creatinine, BUN, bilirubin, total protein, albumin, globulin the albumin:globulin ratio, total cholesterol, hemoglobin A1c, and the urine protein:creatinine ratio. Also, BMI tended to be higher in the positive group than the negative group. While these differences were statistically significant, the numeric differences are quite small with overlapping distributions (see Figure 1) and unlikely to be clinically relevant.

**Table 1:** Characteristics of study population by SARS-CoV-2 antibody status.

|  | **SARS-CoV-2 negative**<br>n = 48,505 | **SARS-CoV-2 positive**<br>n = 1,520 | p |
| --- | --- | --- | --- |
| Age (yrs) | 42 [34,54] | 41 [33,52] | <0.001<sup>2</sup> |
| 18-40 | 47.8% | 50.7% |  |
| 41-60 | 40.2% | 41.4% | 0.55<sup>1</sup> |
| 61-85 | 12.0% | 7.8% |  |
| Sex, (% male) | 55.8% | 53.9% | 0.15<sup>1</sup> |
| Current Smoker | 4.1% | 3.1% | 0.05<sup>1</sup> |
| Heart Disease | 1.2% | 1.0% | 0.61<sup>1</sup> |
| Hypertension | 15.2% | 15.4% | 0.84<sup>1</sup> |
| Diabetes | 4.9% | 4.6% | 0.66<sup>1</sup> |
| Creatinine (mg/dl) | 0.93 [0.79,1.06] | 0.91 [0.77,1.04] | 0.0013<sup>2</sup> |
| BUN (mg/dl) | 14 [11,16] | 13 [11,16] | 0.0031<sup>2</sup> |
| ALKP (IU/L) | 67 [56,82] | 67 [55,81] | 0.599<sup>2</sup> |
| AST (IU/L) | 21 [18,27] | 22 [17,27] | 0.72<sup>2</sup> |
| ALT (IU/L) | 19 [14,28] | 20 [14,29] | 0.307<sup>2</sup> |
| Bilirubin (mg/dl) | 0.47 [0.34,0.65] | 0.44 [0.31,0.62] | <0.001<sup>2</sup> |
| Total Protein (g/dl) | 7.1 [6.8,7.4] | 7.2 [6.9,7.5] | <0.001<sup>2</sup> |
| Albumin (g/dl) | 4.6 [4.4,4.8] | 4.6 [4.4,4.8] | <0.001<sup>2</sup> |
| Globulin (g/dl) | 2.5 [2.2,2.7] | 2.6 [2.3,2.9] | <0.001<sup>2</sup> |
| A:G Ratio | 1.88 [1.67,2.1] | 1.76 [1.54,2] | <0.001<sup>2</sup> |
| Total Cholesterol (mg/dl) | 186 [163,213] | 183 [160,210] | 0.0056<sup>2</sup> |
| HDL (mg/dl) | 56.1 [46.2,67.9] | 55 [46.2,65.7] | 0.014<sup>2</sup> |
| Chol:HDL Ratio | 3.25 [2.67,4.08] | 3.26 [2.7,4.04] | 0.521<sup>2</sup> |
| BMI (kg/m2) | 27.34 [24.21,31.16] | 27.99 [24.74,31.86] | <0.0069<sup>2</sup> |
| Hemoglobin A1c (%) | 5.3 [5.1,5.6] | 5.4 [5.1,5.7] | 0.0069<sup>2</sup> |
| NT-proBNP (ng/ml) | 44 [24,80] | 38 [22,69] | 0.024<sup>2</sup> |
| Urine Protein:Cr Ratio | 0.05 [0.03,0.07] | 0.05 [0.03,0.07] | <0.001<sup>2</sup> |

Numeric values shown as median [IQR]. <sup>1</sup> p-value by Chi-square test. <sup>2</sup> p-value by Mann-Whitney U test.

**Figure 1:**
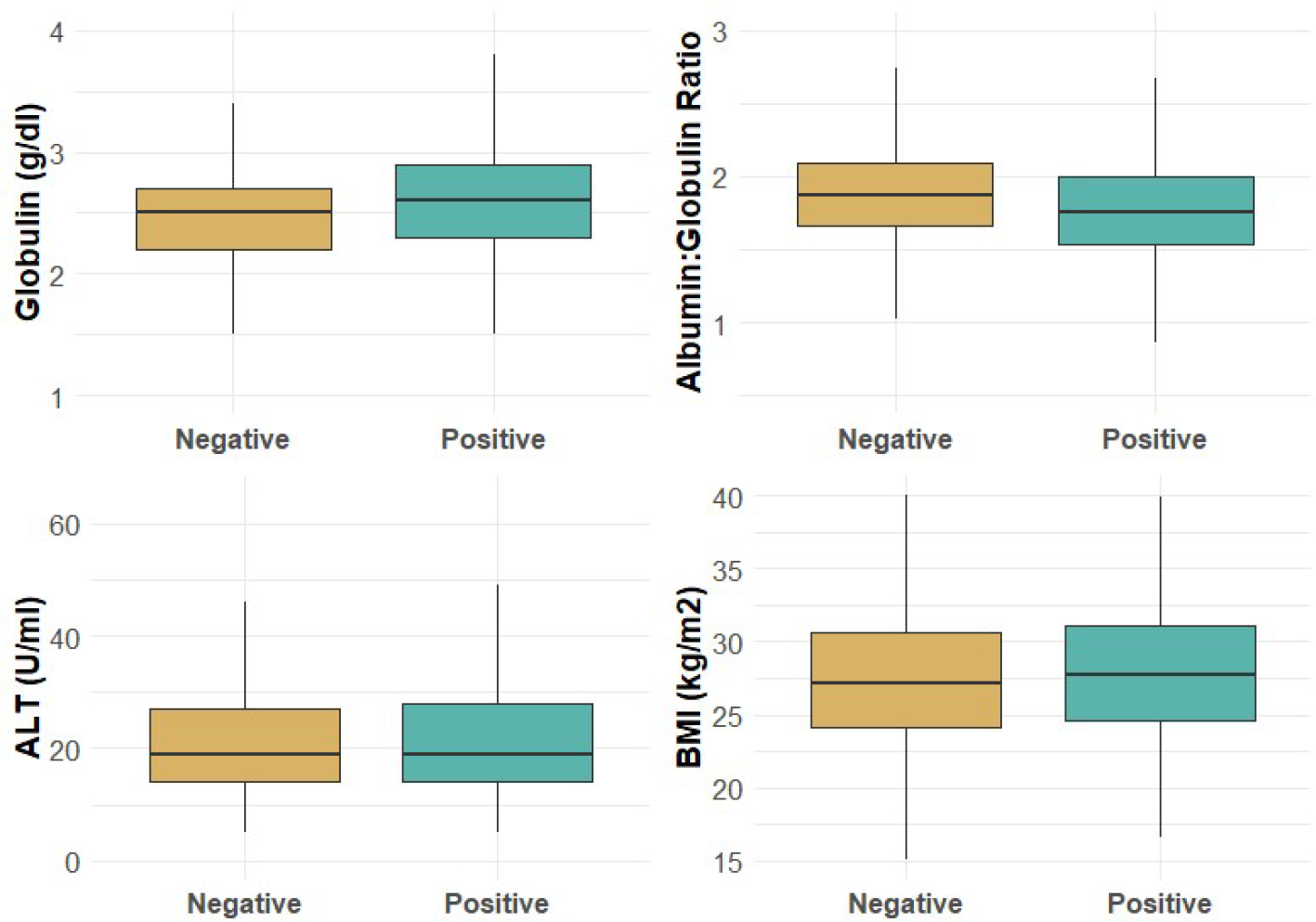
Boxplots of Selected Laboratory Values by SARS-CoV-2 Antibody Status

Rates of positive serology varied by age and sex (see Table 2), with lower rates among individuals over age 60. Geographically, at least 50 samples were obtained from each state and the District of Columbia. Fewer than 100 tests were obtained from Vermont (59), Wyoming (70), DC (78), Maine (81) and Alaska (92). In 3 of these states (Wyoming, Maine and Alaska), no positive cases were detected. The greatest numbers of tests were performed in the states of New York (6560), Texas (4959), and Florida (3828), while the highest rates of seropositivity were seen in New York (17.1%), New Jersey (9.2%) and Connecticut (5.9%) – see figure 2 and Table 3.

**Table 2:** Rate of SARS-CoV-2 Antibodies by Age and Sex

| Age Range | Female | Male |
| --- | --- | --- |
| 20-30 | 3.6% | 3.4% |
| 31-40 | 3.1% | 3.1% |
| 41-50 | 3.7% | 3.0% |
| 51-60 | 3.1% | 2.7% |
| 61-70 | 2.0% | 2.5% |
| 71-80 | 1.0% | 1.1% |

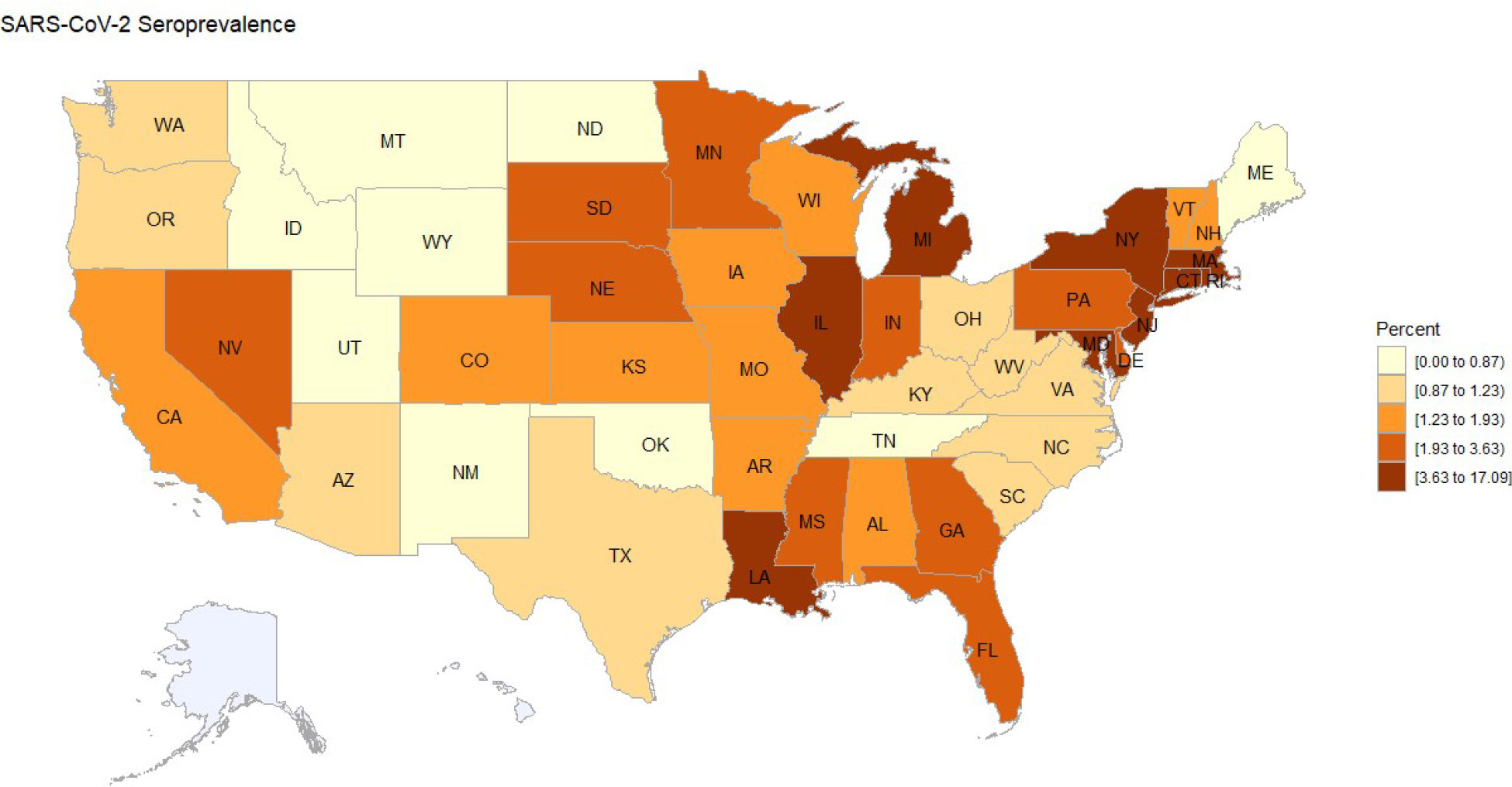
Figure 2:

**Table 3:** Prevalence of SARS-CoV-2 Antibodies by Location

|  |  |  |  |  |  |
| --- | --- | --- | --- | --- | --- |
| **AK** | 0.0% | **KY** | 1.0% | **NY** | 17.1% |
| **AL** | 1.5% | **LA** | 3.8% | **OH** | 0.9% |
| **AR** | 1.4% | **MA** | 4.2% | **OK** | 0.5% |
| **AZ** | 1.1% | **MD** | 4.3% | **OR** | 1.0% |
| **CA** | 1.5% | **ME** | 0.0% | **PA** | 2.6% |
| **CO** | 1.2% | **MI** | 4.4% | **RI** | 4.3% |
| **CT** | 6.0% | **MN** | 2.2% | **SC** | 1.0% |
| **DC** | 5.1% | **MO** | 1.9% | **SD** | 2.2% |
| **DE** | 3.3% | **MS** | 2.2% | **TN** | 0.5% |
| **FL** | 2.0% | **MT** | 0.8% | **TX** | 1.1% |
| **GA** | 2.7% | **NC** | 1.1% | **UT** | 0.7% |
| **HI** | 0.7% | **ND** | 0.8% | **VA** | 1.2% |
| **IA** | 1.5% | **NE** | 2.1% | **VT** | 1.7% |
| **ID** | 0.7% | **NH** | 1.8% | **WA** | 1.1% |
| **IL** | 3.7% | **NJ** | 9.2% | **WI** | 1.3% |
| **IN** | 2.0% | **NM** | 0.5% | **WV** | 0.9% |
| **KS** | 1.6% | **NV** | 2.4% | **WY** | 0.0% |

The estimate of the total number of SARS-CoV-2 infections in the US was 6.98 million (99% CI: 6.56-7.38 million). Because of the small numbers of samples in certain states, the estimation of the overall rate of in the US could not utilize a more precisely weighted approach, but relied solely on state population-based weights. Because antibodies to SARS-CoV-2 may take some time to develop, the total number of COVID reported to the CDC as of June 1^st^, 2020 was used as the baseline comparator. This was chosen because it is in the earlier portion of the date range of the study. Compared to the 1.816 million cases reported to the CDC, our estimate of 6.98 million cases is 3.8 times the total burden of reported cases.

An attempt was made to develop a prediction model (results not shown) using logistic regression and the laboratory values which demonstrated statistically significant differences between the positive and negative groups. These models did not achieve reasonable performance. Even if performance was better it is likely that, over time, as the acuity of the pandemic wanes and antibody levels persist, these lab values will become less predictive of serological status.

## Discussion

This study estimated the seroprevalence of SARS-CoV-2 antibodies in a geographically diverse sample of adults in the US within a 6-week collection period ending in late June 2020. The rate of positivity ranged from 0% to 17% by state and from 1-3% across age and sex categories. The choropleth map of seropositivity roughly corresponds to the areas where the most COVID-19 cases were reported during that period of time. Our results suggest that many more infections occurred than were reported. This is likely due to asymptomatic or minimally symptomatic infections for which care was not sought or symptomatic infection for which testing was not obtained.

Various studies have been published, both before and after peer review, which have reported seroprevalence of SARS-CoV-2 antibodies in the US. Most notably, Havers et al^11^. evaluated a convenience sample (n=16,025) of serological tests on sera submitted to 2 commercial laboratories from 10 US regions. Their estimates of seroprevalence ranged from 1% to 7%, with the highest rates occurring in the New York metro area, Louisiana and Connecticut. The timeframe of this testing differed by region and was earlier than the current study. The authors estimated that the seroprevalence implied that between 6 and 24 times the number of infections had occurred in the studied regions than had been reported.

Stadlbauer et al reported on longitudinal changes in seroprevalence in New York City between late February and mid-April 2020^12^. Over this period of time seroprevalence increased from 2.2% to 10.1%. Rosenberg et al also reported on seroprevalence in the New York metro area^13^. The collection period was from April 19 to 28, 2020, and the estimate was 22.7%. The higher estimate than the current study, despite being performed in an earlier time period, is likely due to a geographical distribution that is more localized to the highest prevalence metro region, rather than the entire state of New York.

Others have studied the relationship between COVID infections, as defined by positive tests for viral nucleic acid sequences, and alterations in standard laboratory tests. There have been no other studies, to our knowledge, reporting relationships between serology results and standard laboratory tests. Joshi et al developed a model to predict PCR positivity^14^. The predictive variables included the absolute neutrophil count (ANC), absolute lymphocyte count (ALC) and the hematocrit. Kurstjens et al developed a similar tool which utilized age, sex, c-reactive peptide, ferritin, lactated dehydrogenase, ALC, ANC and the presence of infiltrates on chest X-ray to predict SARS-CoV-2 PCR positivity, with a final C-statistic of 0.91 in a validation sample^15^.

In the context of COVID-19, standard chemistries and blood count measurements have been studied for their association with, and ability to predict the onset of adult respiratory distress syndrome (ARDS). Wu et al. evaluated those factors associate with ARDS development and found that ANC, LDH, D-dimer, age, hypertension and diabetes were all associated with increased hazard ratios. Jiang et al attempted to develop a prediction algorithm for the development of ARDS. However, it was based on just 53 patients, only 5 of who developed ARDS. They found that ALT was the “most important” predictor but provided no indication of exactly how predictive it was^16^.

The present study is different in that it evaluates laboratory findings in the setting of positive or negative SARS-CoV-2 serology. The difference in lab test results were very modest and insufficient to identify who should or should not be tested for SARS-CoV-2. It implies that, around the time of study, the number of infections in the US was nearly 4 times higher than reported suggesting a much more widespread pandemic, but with a smaller rate of hospitalization, complications and deaths. Weaknesses of the study include the imbalanced representation of the US states, as well as the lack of samples from those under age 20 or over age 80. The age distribution is also more heavily weighted to the young adult years, which is not representative of the US population. Although the sample size was large, it was not large enough to stratify by both age and geography when estimating population seroprevalence. Finally, the life insurance-buying population tends to be both healthier and wealthier than average, and this could also bias the results in an indeterminate direction.

## Conclusion

The rate of SARS-CoV-2 seropositivity in this population of insurance applicants implies a burden of infection approximately 3.8 times higher than the number of reported cases. While some differences in laboratory values reached statistical significance, these differences were numerically small and unlikely to be informative of the probability of testing positive for SARSCoV-2 antibodies.

## Data Availability

Data can be obtained by contacting the authors.

## Declaration of Interest

None

## Notes

### Competing Interest Statement

The authors have declared no competing interest.

### Funding Statement

Funding of the study, including the costs of laboratory testing, were provided by Clinical Reference Laboratories.

### Author Declarations

Western Institutional Review Board (WIRB) IRB Affairs Department reviewed the study under the Common Rule and applicable guidance and determined it is exempt under 45 CFR 46.104(d)(4) using de-identified study samples for epidemiologic investigation.

